# Effectiveness and safety of PRP on persistent olfactory dysfunction related to COVID-19: towards a new therapeutic hope

**DOI:** 10.1101/2022.02.14.22270109

**Authors:** Younès Steffens, Serge Le Bon, Léa Prunier, Alexandra Rodriguez, Jérôme R. Lechien, Sven Saussez, Mihaela Horoi

## Abstract

Olfactory dysfunction (OD) is a well know symptom of coronavirus disease 2019 (COVID-19), accounting for 48 to 85% of patients. In 1 to 10% of cases, patients develop a chronic olfactory dysfunction (COD,) lasting more than 6 months. Recently, platelet-rich plasma (PRP) was used in patients with non-COVID-19 COD and authors reported encouraging results. In the present study, we investigated the usefulness and safety of PRP injection in 56 patients with COVID-19 COD.

Our results showed that PRP in the olfactory cleft can increase the olfactory threshold one month after the injection. Moreover, our results suggest that timing of treatment may be an important factor and that PRP is a safe treatment because no adverse effects were reported throughout the study

## Introduction

Olfactory dysfunction (OD) is a prevalent symptom of coronavirus disease 2019 (COVID-19), accounting for 48 to 85% of patients.^1^ In 1 to 10% of cases, patients develop a chronic olfactory dysfunction (COD,) lasting more than 6 months.^2^ To date, there is no effective therapeutic approach for COD patients. Recently, platelet-rich plasma (PRP) was used in patients with non-COVID-19 COD and authors reported encouraging results.^3^ In the present study, we investigated the usefulness and safety of PRP injection in 56 patients with COVID-19 COD.

## Methods

From January to August 2021, adult patients with COVID-19 COD were prospectively recruited from the University Hospital of Brussels (CHU Saint-Pierre, Belgium). The COVID-19 diagnosis was based on positive RT-PCR findings. Patients with abnormal findings on nasal endoscopy (e.g., polyposis, rhinosinusitis); blood disorders, or blood thinner use were excluded. Participants benefited from olfactory testing (Sniffin’ Sticks Test (Medisense, Groningen, Netherlands) resulting in the Threshold Discrimination Identification TDI score) at baseline and 1 month post-injection. The improvement of olfactory function was evaluated with a Likert-scale ranging from 0 (none) to 3 (strong).

PRP injections were performed in each olfactory cleft via nasal endoscopy and under local anesthesia by the same physician (YS), following the protocol of Yan *et al*. (GS30-PURE II Protocol A: Emcyte, Ft Myers, Florida).^3^ According to the data distribution, the following tests were used: Wilcoxon signed-rank test, Mann–Whitney test, and Spearman correlation test. A level of significance of p<.05 was used.

We compare the result of the PRP group to a control group how underwent simple olfactive training for one month.

## Results

Thirty-six patients received PRP injection. Among those, six were lost to follow-up and therefore excluded. The control group matched for age, gender and TDI score at baseline included 26 patients with COVID-19 COD. Both groups were comparable regarding demographics, duration of OD, and TDI (**Table**). At 1 month post-PRP injection, the mean TDI score significantly improved by 6.7 points in the PRP group (p<.001; **Figure**), while there was no significant change in controls. There was a moderate negative correlation between TDI score difference and duration of OD in the PRP group (r=.387, p=.035) but not in controls. The mean self-assessment of improvement in smell function was 1.8 (mild-to-moderate) in the PRP group, which was significantly higher than the score (0.3) of healthy individuals (p<.001). No adverse effects were reported throughout the study.

**Figure 1.**
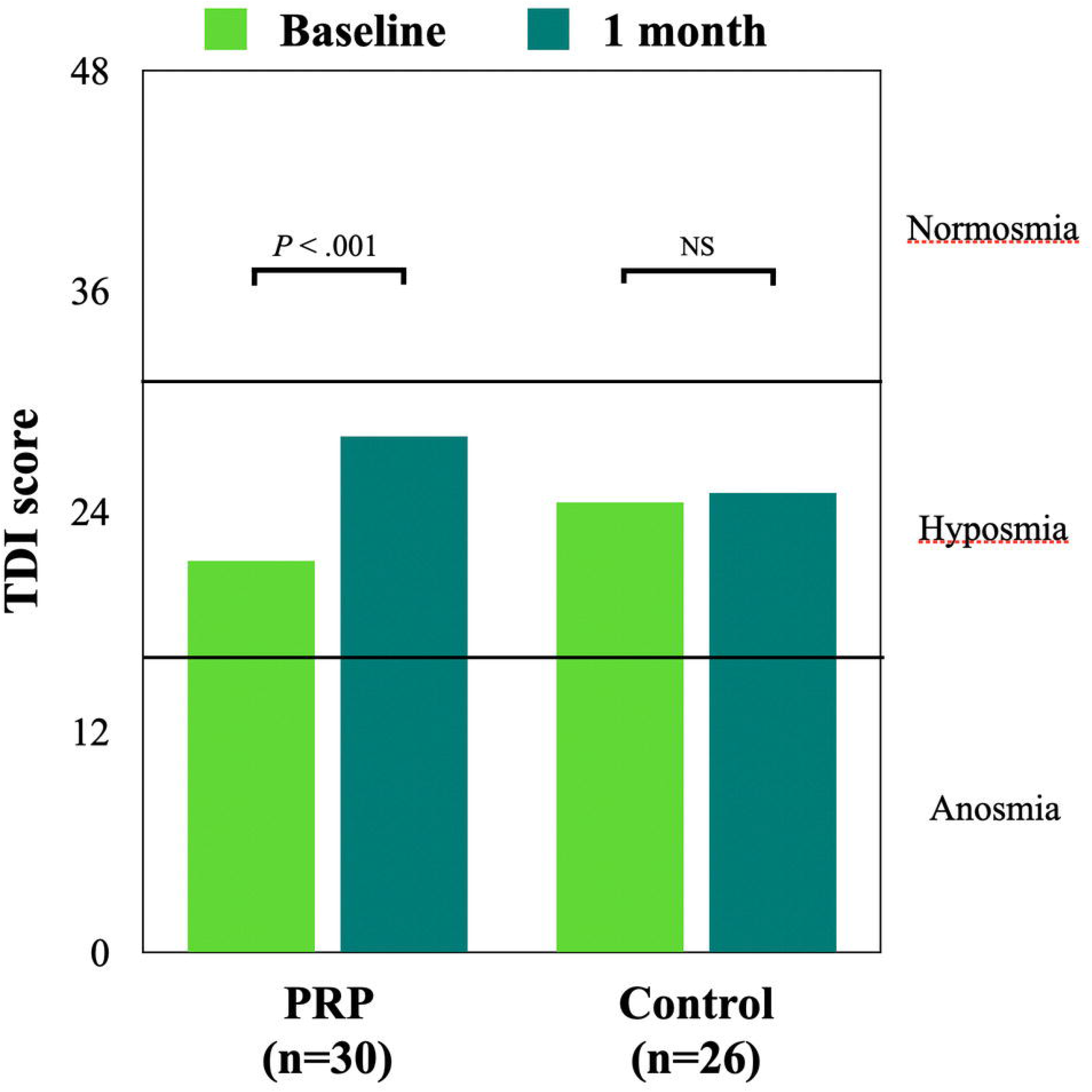
Evolution of TDI score at baseline and after one month in both PRP and control groups. Abbreviations: NS, not significant; PRP, platelet-rich plasma; TDI, Threshold-Discrimination-Identification

**Table 1.** Comparison of demographics, olfactory and subjective scores between PRP and control groups.

|  | PRP group<br>(n = 30) | Control group<br>(n = 26) | p-value |
| --- | --- | --- | --- |
| Age (mean± SD), years | 39 ± 12 | 44 ± 11 | NS |
| Sex (F/M) | 16/14 | 20/6 | NS |
| Duration of olfactory disorder (mean± SD; min - max), months | 10.8 ± 2.5<br>7 - 16 | 9.7 ± 3.4<br>6-17 | NS |
| TDI at baseline (mean± SD) | 21.3 ± 7.4 | 24.5 ± 7.4 | NS |
| TDI at 1 month (mean± SD) | 28.0 ± 5.0 | 25.0 ± 7.7 | NS |
| Subjective improvement (mean± SD) | 1.8 ± 1.0 | 0.3 ± 0.6 | <0.001 |

Abbreviations: NS, not significant; SD, standard deviation; TDI, Threshold-Discrimination-Identification

When we separate the patient considering the duration of olfactory loss, we did not find significant difference (p=0,093) between the group with the loss of smell since less than 12 months and those since more than 12 months.

The average TDI gain at one month was 9 points in the 6–12month group compared to 4 in the 12-18month group.

## Discussion

The main finding of the present preliminary study is the safety and potential usefulness of PRP as an in-office approach improving smell function in psychophysical tests and subjective assessment. One month after PRP injection, mean olfactory score increased by 6.7 points, above the minimal clinically important difference of 5.5-6 for TDI.^4^

In a recent study, De Melo GD et al^5^ suggest that protracted viral infection and inflammation in the olfactory neuroepithelium may account for COVID-19 COD.

This might result from direct damage to the olfactory neuro-epithelium.

PRP is an autologous biologic product derived from fresh whole blood containing a high concentration of platelets. PRP is reported to have anti-inflammatory and pro-regenerative properties, including upregulation of growth factors and neuro-regeneration.^6,7^ Our results corroborate the findings of Yan *et al*. who reported a significant improvement in TDI score one month after PRP injection in the olfactory cleft of seven patients with non-COVID-19 COD.^3^ The high concentration of growth factors in alpha granules in PRP, such as EGF and PDGF, has been found to enhance epithelial and neuro-regeneration.^6,7^

## Conclusion

Our results support that PRP may be especially appropriate in olfactory cleft where both olfactory neuro-epithelium and sustentacular cells have been shown to be injured by the virus. Moreover, our results suggest that timing of treatment may be an important factor: patients with earlier PRP injection showed higher olfactory score change than patients with a longer duration of olfactory disorder. Future randomized controlled studies are needed to confirm these results and explore long-term effect of this new approach.

## Data Availability

All data produced in the present study are available upon reasonable request to the authors

## Notes

### Competing Interest Statement

The authors have declared no competing interest.

### Clinical Trial

NCT05226546

### Funding Statement

This study did not receive any funding

### Author Declarations

This study was approved by the Institutional Review Board (CHUSP2102028)

